# Pilot Study of Large Language Models as an Age-Appropriate Explanatory Tool for Chronic Pediatric Conditions

**DOI:** 10.1101/2024.08.06.24311544

**Authors:** Cameron C. Young, Elizabeth Enichen, Arya Rao, Sidney Hilker, Alex Butler, Jessica Laird-Gion, Marc D. Succi

## Abstract

There exists a gap in existing patient education resources for children with chronic conditions. This pilot study assesses large language models’ (LLMs) capacity to deliver developmentally appropriate explanations of chronic conditions to pediatric patients. Two commonly used LLMs generated responses that accurately, appropriately, and effectively communicate complex medical information, making them a potentially valuable tool for enhancing patient understanding and engagement in clinical settings.

## Introduction

The ability to translate complex medical terminology into commonly understood phrases is one of the numerous promising applications of artificial intelligence (AI), particularly large language models (LLMs), in the healthcare field.^1-8^ LLMs are advanced AI models designed to understand and generate human-like text by leveraging vast amounts of data and complex algorithms. Communicating medical information to children with chronic conditions presents a unique challenge for providers as developmental stages, perspectives, and understanding vary considerably across ages and disease processes.^9^ Previous studies have shown that how providers communicate can affect both health outcomes and patient and caregiver satisfaction;^10,11^ particularly, ineffective communication can result in negative outcomes for children and families.^12,13^ Therefore, ensuring children comprehend health information empowers active participation in their medical care, increasing knowledge and treatment adherence, while reducing adverse events.^14,15^

There exists a gap in educational materials for pediatric patients with chronic conditions due to the lack of standardized approaches, particularly for rare diseases, indicating a scarcity of research in this area. Current materials often fail to cater to the specific needs of pediatric patients, neither being written in age-appropriate, plain language nor considering the complexities of multisystemic diseases, or focus on educating the parents, rather than the patient.^15^ Recent studies emphasize the significance of tailoring educational programs to meet the unique needs of pediatric patients with chronic conditions. For instance, a component-based educational program was successful in improving self-efficacy and treatment satisfaction among children with rare chronic diseases.^16^

LLMs offer a novel solution to this challenge. Given this potential, we hypothesize that LLMs can serve as effective tools for providing age-appropriate explanations of chronic conditions, thereby enhancing the communication between healthcare providers, caregivers, and pediatric patients. This study evaluates the ability of two commonly used LLMs to generate accurate, complete, and developmentally appropriate explanations of chronic diseases to children of different ages. By integrating these AI tools into pediatric healthcare communication, we aim to bridge the gap between clinical knowledge and patient comprehension, fostering better engagement and adherence to treatment among young patients.

## Methods

Two generalist LLMs (GPT-4 [OpenAI] and Gemini 1.0 Ultra [Google]; accessed January 16, 2024) were asked to respond to the following prompt: “act as a pediatrician and explain a diagnosis of [CONDITION] to a [AGE]-year-old in language they can understand.” Responses were generated for five common chronic conditions (asthma, anaphylactic allergy [peanut allergy], epilepsy, sickle cell disease, and type I diabetes) for children of odd ages between 5 and 17 (5-year-old, 7-year-old, 9-year-old, 11-year-old, 13-year-old, 15-year-old, and 17-year-old). Representative responses from GPT-4 and Gemini can be found in **Supplementary Table 1**.

A total of 70 LLM responses (35 from each model) were assessed for accuracy, completeness, age-appropriateness, possibility of demographic bias, and overall quality, based on an existing framework for the human evaluations of the clinical application of LLMs and prior literature.^17^ Demographic bias was defined as whether implementing the response in clinical practice would favor or disadvantage particular groups based on demographic characteristics such as race, age, gender, socioeconomic status, or geographic location. Three pediatric physicians (S.H., A.B., and J.L.) rated the responses based on how well they aligned with these five criteria using a Likert scale from 1 (highly disagree) to 5 (highly agree). Numeric ratings were treated as continuous variables and summarized as means and 95% confidence intervals. A Welch two sample t-test was used to assess differences in means. P<0.05 was considered statistically significant. Intra-rater reliability was assessed by calculating Pearson correlation coefficients between individual raters. Additionally, Pearson correlation coefficients were computed to assess the degree of correlation between evaluation criteria Analyses were performed in R version 4.2.2.

## Results

Across both LLMs, responses were rated as highly accurate (GPT-4: 4.37 [4.27-4.47]; Gemini: 4.55 [4.45-4.65]), highly complete (GPT-4: 4.25, [4.16-4.34]; Gemini: 4.39, [4.28-4.50]), moderately age-appropriate (GPT-4: 3.95, [3.81-4.09]; Gemini: 3.26, [3.09-3.43]), of moderate quality (GPT-4: 3.88, [3.75-4.01]; Gemini: 3.43, [3.26-3.60]), and with low possibility of demographic bias (GPT-4: 1.61, [1.49-1.73]; Gemini: 1.16, [1.11-1.21]). Gemini responses had a significantly lower possibility of demographic bias (p<0.001), while responses from GPT-4 were of significantly higher quality (p=0.004) and age-appropriateness (p<0.001) (**Table 1**). Across both models, age-appropriateness and overall quality tended to increase with age, while other criteria remained similar (**Table 2**). There were no differences in ratings across chronic conditions (**Supplementary Table 2**). Intra-rater reliability was high, with an average Pearson correlation coefficient of 0.72 (**Supplementary Table 3**).

**Table 1.** Overall and age-stratified average reviewer ratings of GPT-4 and Gemini across five evaluation criteria.

| **Large Language Model** | **Accuracy, mean (95% CI)** | **Completeness, mean (95% CI)** | **Age-Appropriateness, mean (95% CI)** | **Possibility of Demographic Bias, mean (95% CI)** | **Overall Quality, mean (95% CI)** |
| --- | --- | --- | --- | --- | --- |
| *GPT-4* | 4.37 (4.27, 4.47) | 4.25 (4.16, 4.34) | 3.95 (3.81, 4.09) | 1.61 (1.49, 1.73) | 3.88 (3.75, 4.01) |
| *Gemini* | 4.55 (4.45, 4.65) | 4.39 (4.28, 4.50) | 3.26 (3.09, 3.43) | 1.16 (1.11, 1.21) | 3.43 (3.26, 3.60) |
| *P-value* | 0.08 | 0.15 | **<0.001** | **<0.001** | **0.004** |

CI = confidence interval

**Table 2.** Age-stratified average reviewer ratings of GPT-4 and Gemini responses across five evaluation criteria.

| **Large Language Model** | **Accuracy, mean (95% CI)** | **Completeness, mean (95% CI)** | **Age-Appropriateness, mean (95% CI)** | **Possibility of Demographic Bias, mean (95% CI)** | **Overall Quality, mean (95% CI)** |
| --- | --- | --- | --- | --- | --- |
| *GPT-4* |  |  |  |  |  |
| *5-Year-Old* | 4.20 (3.76, 4.64) | 4.07 (3.67, 4.47) | 3.47 (2.76, 4.18) | 1.53 (1.07, 1.99) | 3.47 (2.87, 4.07) |
| *7-Year-Old* | 4.40 (4.08, 4.72) | 4.20 (3.99, 4.41) | 4.07 (3.62, 4.52) | 1.53 (1.15, 1.91) | 3.93 (3.63, 4.23) |
| *9-Year-Old* | 4.47 (4.21, 4.73) | 4.27 (3.97, 4.57) | 4.07 (3.71, 4.43) | 1.60 (1.28, 1.92) | 3.93 (3.57, 4.29) |
| *11-Year-Old* | 4.40 (3.94, 4.86) | 4.27 (3.97, 4.57) | 4.00 (3.57, 4.43) | 1.33 (1.08, 1.58) | 3.80 (3.32, 4.28) |
| *13-Year-Old* | 4.27 (3.91, 4.63) | 4.13 (3.75, 4.51) | 3.87 (3.33, 4.41) | 1.73 (1.24, 2.22) | 3.93 (3.41, 4.45) |
| *15-Year-Old* | 4.40 (3.98, 4.82) | 4.40 (4.08, 4.72) | 3.67 (2.91, 4.43) | 1.93 (1.34, 2.52) | 3.93 (3.31, 4.55) |
| *17-Year-Old* | 4.47 (4.09, 4.85) | 4.40 (4.08, 4.72) | 4.53 (4.27, 4.79) | 1.60 (1.07, 2.13) | 4.13 (3.81, 4.45) |
| *Gemini* |  |  |  |  |  |
| *5-Year-Old* | 4.47 (4.01, 4.93) | 4.27 (3.82, 4.72) | 2.53 (1.79, 3.27) | 1.33 (1.02, 1.64) | 2.87 (2.18, 3.56) |
| *7-Year-Old* | 4.53 (4.11, 4.95) | 4.40 (3.98, 4.82) | 2.53 (1.90, 3.16) | 1.07 (0.94, 1.20) | 3.07 (2.32, 3.82) |
| *9-Year-Old* | 4.60 (4.14, 5.06) | 4.47 (4.09, 4.85) | 3.00 (2.37, 3.63) | 1.20 (0.99, 1.41) | 3.20 (2.51, 3.89) |
| *11-Year-Old* | 4.60 (4.28, 4.92) | 4.40 (4.03, 4.77) | 3.00 (2.49, 3.51) | 1.07 (0.94, 1.20) | 3.07 (2.48, 3.66) |
| *13-Year-Old* | 4.67 (4.42, 4.92) | 4.27 (3.91, 4.63) | 3.80 (3.32, 4.28) | 1.13 (0.95, 1.31) | 4.00 (3.57, 4.43) |
| *15-Year-Old* | 4.60 (4.23, 4.97) | 4.47 (4.01, 4.93) | 3.80 (3.52, 4.08) | 1.20 (0.99, 1.41) | 3.87 (3.41, 4.33) |
| *17-Year-Old* | 4.47 (4.01, 4.93) | 4.27 (3.82, 4.72) | 2.53 (1.79, 3.27) | 1.33 (1.02, 1.64) | 2.87 (2.18, 3.56) |

CI = confidence interval

The use of metaphors to explain biological concepts was common throughout responses (red blood cells are “delivery trucks” around the body, insulin is the “key” to unlocking the door for glucose to enter cells, a “glitch” in the brain causes an epileptic seizure). References to superheroes (15.7% of responses), food (12.9% of responses), and weather (12.9% of responses) were most frequent among all responses. Additionally, the mention of videogames, sports, and cartoons were common. Some of these responses were confusing in the context that they were provided (“villains blocking pipes” in a videogame may not be easily understandable by all children), could be interpreted as problematic by the patient (a “glitch in the brain” may seem that something is wrong that can never be fixed), or risk demographic bias (referring to a child as “kiddo” or “buddy”).

## Discussion

LLMs can generate accurate, complete, age-appropriate chronic disease explanations with low possibility of demographic bias for children of different ages and chronic conditions, providing a potential additional source of patient educational materials. These models are flexible, easy-to-use, and can be implemented at the point of care by clinicians or at home by parents or caregivers and personalized to a patient’s specific condition and demographics. Further, technology-based interventions can positively impact pediatric health-related outcomes,^18^ further highlighting the potential utility of these tools.

Additionally, the use of AI chatbots is popular among children and adolescents through their integration into social media platforms, such as Snapchat’s My AI^19^ and as educational tools.^20^ Further, a survey of parents showed an openness towards AI-driven technologies in pediatric healthcare, with quality, convenience, and cost positively influencing their openness, but concerns about privacy, the need for human interaction in care, and shared decision-making were noted.^21^

Despite these positive findings and likelihood of translatability, there are several limitations related to the findings. The use of words like “kiddo” or “buddy” as well as references to sports and videogames may risk biasing patients and decreasing effectiveness of explanations.^14^ Further, differences in age-appropriateness, possibility of demographic bias, and overall quality were noted between GPT-4 and Gemini. This discrepancy in LLM responses could be due to variations in training data and model architecture.^22^ Therefore, clinicians should be cognizant of these potential differences, and evaluate multiple LLM output before sharing responses with patients and caregivers. Finally, these responses were reviewed by pediatric clinicians, rather than children, who may interpret these responses differently. Evaluation of children’s interactions with LLMs for pediatric healthcare represents a promising area of future research.

This pilot study shows that LLMs offer a promising tool to explain complex chronic diseases to children of different ages, with room for improvement. Developing custom-built, specialty LLMs curated by clinicians and child development experts that incorporate patient-specific details may improve these LLMs ability to act as an explanatory tool.^9^ However, LLMs have the potential to aid in closing the existing gap in education materials for pediatric patients with chronic conditions.

## Data Availability

All data will be made available for any research purpose by contacting the corresponding author.

## Notes

**Conflicts of Interest:** The authors have no relevant financial or non-financial interests to disclose.

### Competing Interest Statement

The authors have declared no competing interest.

### Funding Statement

The project described was supported in part by award Number T32GM144273 from the National Institute of General Medical Sciences. The content is solely the responsibility of the authors and does not necessarily represent the official views of the National Institute of General Medical Sciences or the National Institutes of Health.

