## Supplemental Material for "Pilot Study of Large Language Models as an Age-Appropriate Explanatory Tool for Chronic Pediatric Conditions"

**Table S1** – Representative responses from large language models

|  | **GPT-4** | **Gemini** |
| --- | --- | --- |
| 13-year-old with asthma | Hey there, buddy! So, you know how when you're playing a video game and sometimes your character runs out of energy and needs to recharge? Well, our bodies are kind of like that too. We have these tubes in our bodies called bronchial tubes that help us breathe by carrying air in and out of our lungs.  Now, imagine these tubes are like the pipes in a video game or a cartoon. When you have asthma, these tubes can sometimes get a little smaller and tighter, kind of like when a villain in a game puts a blockage in the pipe. This makes it harder for the air to move in and out, and that's why you might feel like you're running out of breath or your chest feels tight.  Also, sometimes, these tubes can get a little irritated and produce more mucus. Think of it like the slime monsters in your games. This extra mucus can make the tubes even narrower, making it even harder to breathe.  But don't worry, we have power-ups in real life too! There are medicines that can help open up these tubes again, making it easier for you to breathe. Some medicines you take every day to keep the tubes open, and some you take when you feel like you're having trouble breathing.  Just like in a game, it's important to know when to use your power-ups and to avoid things that make your asthma worse, like dust or smoke. And remember, you can always talk to me or your parents if you're feeling unwell or have any questions. | Asthma is a condition that makes your airways, which are the tubes that carry air to your lungs, become inflamed and narrow. When this happens, it can make it harder for you to breathe. That's why you've been having shortness of breath, coughing, and wheezing.  Think of your airways like straws. In someone without asthma, the straws are nice and wide open, allowing air to flow in and out easily. But in someone with asthma, the straws are more like narrow, bendy hoses. They can get squeezed and blocked easily, making it hard to breathe.  The good news is that asthma is a very treatable condition. We have lots of medications that can help keep your airways open and prevent asthma attacks. We also have some great tools that can help you manage your asthma, like peak flow meters and spacers.  With the right treatment and management, you can live a normal, active life with asthma. You'll be able to do all the things you love to do, like play sports, go to school, and hang out with your friends.  Here are a few things you can do to help manage your asthma:   - Take your medications as prescribed. This is the most important thing you can do to keep your asthma under control. - Use a peak flow meter to monitor your asthma. This device will help you measure how well your lungs are working. - Avoid things that trigger your asthma, such as smoke, dust, and pollen. - Wash your hands often to prevent colds and flu, which can make asthma worse. - Get regular exercise. Exercise can help strengthen your lungs and improve your overall health.   I know it can be scary to have asthma, but I want to assure you that it is a very common condition. Millions of kids and adults have asthma, and we have lots of resources to help you manage it. With the right treatment and care, you can live a full and happy life with asthma. |
| 7-year-old with type I diabetes | Hey kiddo, you know how we eat food every day? Well, our body uses that food to make energy so we can run, play, and do all the things we love. To do this, our body needs a helper called insulin. Insulin is like a key that opens the door to our body's cells so that sugar from the food can enter and be used for energy.  Now, in your body, the place that makes insulin, called the pancreas, is having a little trouble. It's not making enough insulin. This means that the sugar from the food you eat can't get into your cells to give you energy. Instead, it stays in your blood, and that's not good.  This is called Type 1 Diabetes. It's not because of anything you did, and it's not something you can catch like a cold. It just happens to some people. But don't worry, we have ways to help your body get the insulin it needs. You'll need to take insulin every day, and we'll teach you and your parents how to do that. You'll also need to check your blood sugar levels regularly to make sure they're not too high or too low.  It might seem a bit scary now, but with some practice, it will become a normal part of your day, just like brushing your teeth. And remember, you can still do all the things you love. You just have to take a few extra steps to stay healthy. | Hi there! I see you are here because you are experiencing some changes in your body. That's because you have diabetes.  Diabetes is a condition where your body has trouble using a type of sugar called glucose for energy. Normally, the pancreas makes insulin, a special chemical that helps glucose get from your blood into your cells. But in people with diabetes, the pancreas doesn't make enough insulin or doesn't make any at all.  Without insulin, glucose builds up in your blood instead of being used for energy. High glucose levels can make you feel tired, thirsty, and have to use the bathroom a lot. It can also make it hard for your body to heal cuts and infections.  The type of diabetes you have is called type 1. This means that your pancreas doesn't make any insulin. People with type 1 diabetes need to take insulin every day to stay healthy.  Taking insulin is like giving your body the key to unlock the door so glucose can get into your cells. You can take insulin in shots or through a special pump that goes under your skin.  I know this all may seem a little scary at first, but I want to assure you that diabetes is something that you can manage. With the right treatment, you can live a long, healthy, and happy life. We will work together to make sure you get the care you need.  Having diabetes means that you need to be extra careful to take care of yourself. That means:   - Eating healthy foods - Getting regular exercise - Monitoring your blood sugar levels - Taking your insulin as prescribed   I know it may be hard to remember all of these things at first, but we will work together to create a plan that works for you. And I promise, it will get easier over time.  Just remember, you are not alone. There are millions of people around the world who have diabetes. And I'm here to help you every step of the way. |

**Table S2** – Disease-stratified average reviewer ratings of GPT-4 and Gemini across five evaluation criteria

| **Large Language Model** | **Accuracy, mean (95% CI)** | **Completeness, mean (95% CI)** | **Age-Appropriateness, mean (95% CI)** | **Possibility of Demographic Bias, mean (95% CI)** | **Overall Quality, mean (95% CI)** |
| --- | --- | --- | --- | --- | --- |
| *GPT-4* | | | | | |
| Asthma | 4.24 (4.00, 4.48) | 4.19 (3.90, 4.48) | 3.57 (3.11, 4.03) | 1.81 (1.66, 1.96) | 3.76 (3.26, 4.26) |
| Anaphylaxis | 4.33 (4.20, 4.46) | 4.29 (4.09, 4.49) | 4.05 (3.49, 4.61) | 1.90 (1.75, 2.05) | 3.95 (3.47, 4.43) |
| Epilepsy | 4.33 (3.94, 4.72) | 4.10 (3.75, 4.45) | 4.38 (3.83, 4.93) | 1.90 (1.71, 2.09) | 4.10 (3.50, 4.70) |
| Sickle Cell Disease | 4.52 (4.23, 4.81) | 4.33 (4.01, 4.65) | 4.10 (3.57, 4.63) | 1.10 (1.01, 1.19) | 4.05 (3.52, 4.58) |
| Type 1 Diabetes | 4.43 (3.98, 4.88) | 4.33 (3.91, 4.75) | 3.67 (3.10, 4.24) | 1.33 (1.10, 1.56) | 3.52 (2.96, 4.08) |
| *Gemini* | | | | | |
| Asthma | 4.71 (4.47, 4.95) | 4.48 (4.19, 4.77) | 3.48 (3.02, 3.94) | 1.14 (0.99, 1.29) | 3.76 (3.26, 4.26) |
| Anaphylaxis | 4.90 (4.77, 5.03) | 4.71 (4.51, 4.91) | 3.24 (2.68, 3.8) | 1.14 (0.99, 1.29) | 3.48 (3.00, 3.96) |
| Epilepsy | 4.33 (3.94, 4.72) | 4.43 (4.08, 4.78) | 3.05 (2.50, 3.60) | 1.24 (1.05, 1.43) | 3.43 (2.83, 4.03) |
| Sickle Cell Disease | 4.57 (4.28, 4.86) | 4.38 (4.06, 4.70) | 3.33 (2.8, 3.86) | 1.05 (0.96, 1.14) | 3.38 (2.85, 3.91) |
| Type 1 Diabetes | 4.24 (3.79, 4.69) | 3.95 (3.53, 4.37) | 3.19 (2.62, 3.76) | 1.24 (1.01, 1.47) | 3.10 (2.54, 3.66) |

**Table S3** – Pearson correlation coefficients between reviewer ratings

| **Reviewer Combination** | **Pearson Correlation Coefficient** |
| --- | --- |
| *Reviewer 1/2* | 0.771 |
| *Reviewer 1/3* | 0.661 |
| *Reviewer 2/3* | 0.719 |

**Figure S1** – Pearson correlation coefficient matrix of evaluation criteria

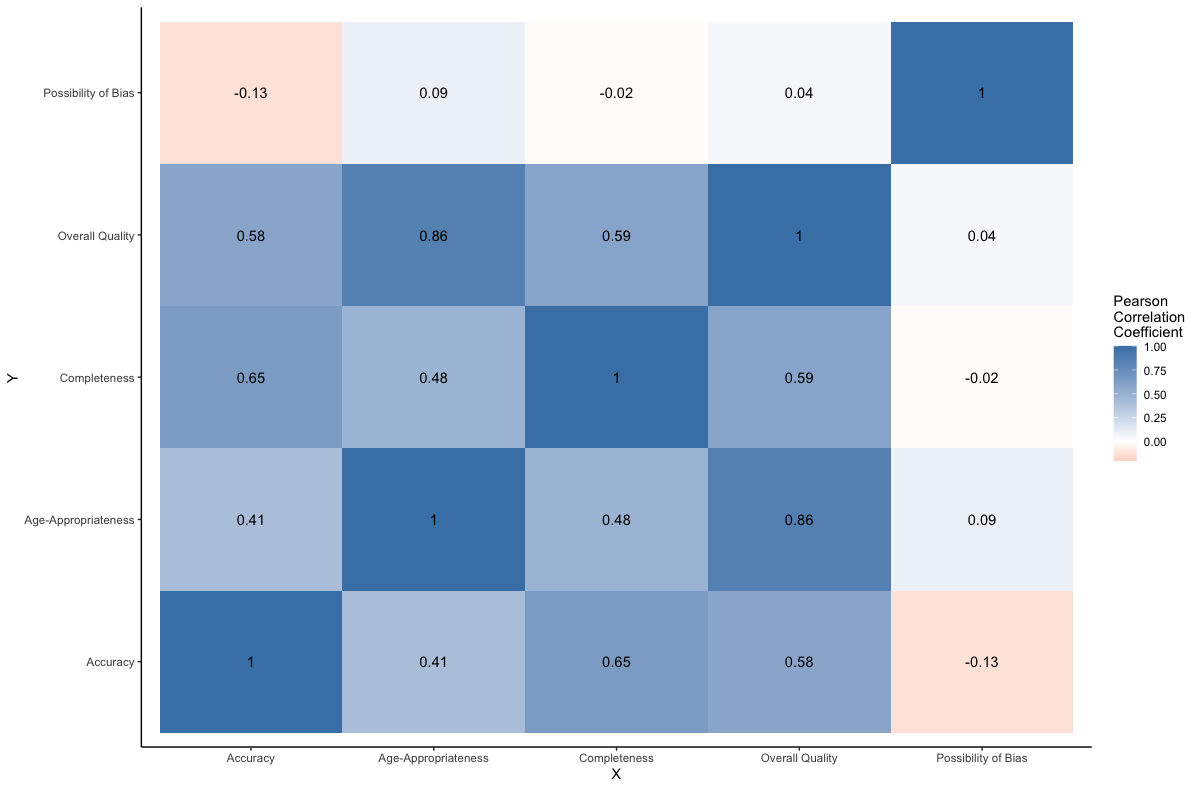
